# Omeprazole and cardiovascular risk via induction of statin dysmetabolism

**DOI:** 10.1101/2024.10.14.24315493

**Authors:** Eugene Chen Howe Goh, Lik Hang Wu, Leroy Pakkiri, Mei Li Ng, Maya George, JL Yang, XY Liu, Folefac Aminkeng, K Muthu Shanmugam, Jian Jun Liu, R Roshni Singaraja, A Elisa Liehn, E Shyong Tai, Jack Tan, Chester Lee Drum

## Abstract

**Background:** Statins are a first line treatment for reducing cardiovascular risk. While drug-drug interactions which cause symptomatic side effects are well known, drug-drug interactions which decrease statin effectiveness are poorly understood.

**Methods:** A multicentre prospective observational study of 1362 participants on chronic statin medication underwent quantitative mass spectrometry (LC-MS/MS) of statin metabolites up to 13 hours post-dose. Participants were followed up to 5 years for major adverse cardiovascular events (MACE) as a composite of stroke, acute myocardial infarction and all-cause mortality. Linear modelling of statin lactone with co-prescribed medications, and multivariable cox proportional hazards modelling of statin metabolites, co-prescriptions and 5-year MACE were performed. An interventional mouse model of atheroma progression was used to validate the results.

**Results:** There were 1139 (83.6%) male and 223 (16.4%) female participants, mean age was 59±9.4. Of all 51 co-prescribed drugs, omeprazole was the second most common (57.4%), and was the strongest determinant of increased plasma statin lactone, demonstrating a ∼1.41 fold increase in lactone concentration and a 1.46-fold increase in 5-year rates of major adverse cardiovascular events (MACE). Independent of statin drug dose and achieved LDL cholesterol levels, plasma atorvastatin lactone (ATVLAC) levels ≥3.9ng/mL strongly predicted MACE (HR=2.45) and all-cause mortality (HR=3.18). In a mouse model of atheroma formation, omeprazole co-administration with atorvastatin resulted in increased lactonization (1.87-fold) and significantly higher lesion area (2.97-fold).

**Conclusion:** Omeprazole co-prescription with atorvastatin represents a novel drug-drug interaction mediated by increased statin lactonization and is strongly associated with increased 5-year MACE.

**Registration:** URL: https://clinicaltrials.gov/; ID: NCT03042286

**Clinical Perspective:** *What is New?:* Discovery of novel drug-drug interactions are difficult because drug metabolism is not directly measured in clinical care. In this study, we used a well powered prospective study to quantitatively phenotype statin metabolism and discover medical and clinical determinants of dysmetabolism. Co-prescription of omeprazole was strongly associated with statin lactone production (a statin metabolite that does not affect HMG-CoA reductase and is postulated to cause off target side effects). These patients had higher rates of MACE, independent of classical predictors such as LDL cholesterol and other Framingham risk factors. In mice co-administered either omeprazole or lansoprazole with statin, both statin lactonization and neointimal plaque formation were increased.

*What are the clinical implications?:* This study provides insights into a novel drug-drug interaction between statins and proton pump inhibitors, two of the most prescribed medication classes in the world, suggesting that this class of drugs should not be prescribed with statins. We also demonstrate a novel methodology for discovering DDI’s through the direct quantification of drug metabolism in real-world clinical cohorts. The findings may be of use in precision medicine initiatives regarding cardiovascular management and statin use.

## Main Text

Statins are an effective treatment for the reduction of cardiovascular risk and are the second most-common class of prescribed medicines, with ∼200 million users worldwide.^1^ Proton pump inhibitors (PPI’s), reduce acid secretion in the stomach and are the most common class of medicines prescribed globally, estimated to be taken by ∼23.4% of the adult population in the developed world^2^. In the setting of cardiovascular risk, PPI’s are commonly prescribed to reduce rates of gastrointestinal bleeding. Observational studies consistently associate PPI usage with adverse cardiovascular outcomes, raising concerns for undiscovered risk factors and possible over prescription of this drug class^3,4^.

Statins benefit patients through the inhibition of HMG-CoA Reductase and subsequent lowering of LDL cholesterol, mediated by the acid form of the drug. Statins also have known side effects, including symptomatic muscle toxicity, which are attributed to a hydrophobic metabolite of the drug, characterized by a lactone ring replacing the acidic dihydroxyheptanoic structure of the drug. The carboxy moiety is involved in the inhibition of HMG-CoA reductase. Thus the lactone form plays no role in cholesterol reduction. Instead, statin lactones permeate cellular membranes in an unregulated fashion and potently inhibit mitochondrial function^5^.

PPI’s inhibit H+/K+ ATPase, the enzyme responsible for creating the acidic environment of the stomach, and are prescribed to treat symptoms of gastroesophageal reflux (GERD). PPI’s also reduce rates of gastrointestinal bleeding and are thus commonly prescribed to cardiovascular patients under intensive medical management, which includes statins, antihypertensives and concomitant antiplatelet or anticoagulation therapy. Despite this, PPI’s are regularly associated with adverse cardiac outcomes, with hazard ratios for MACE ranging from 1.21 to 1.37^6^.

Statins and PPI’s are two of the most widely prescribed medicines in the world. No DDI’s to date have been reported between the drug classes, however a theoretical basis for PPI-statin DDI exists with reference to the PPI based induction of *UGT1A*^7^, an enzyme which is responsible for lactonization of statin acids in humans. Because of the common use of statins in the setting of cardiovascular risk, we hypothesized that pharmacological and genetic influences on statin lactonization may have an adverse effect on cardiovascular outcomes. In this study we used a combination of mass spectrometry and clinical phenotyping, combined with 5-year outcomes analyses, to investigate determinants of lactonization and cardiovascular outcome in individuals using statin.

## Methods

For additional details on all methods, please refer to Supplementary Note.

### Experimental Model and Subject Details

#### Human Study Design

This was a prospective observational multicentre cohort study (SAPhIRE, NCT03042286) that began in 2016 and conducted at 2 independent sites in Singapore, the National University Hospital (NUH) and the National Heart Centre, Singapore General Hospital (SGH), consisting of patients on chronic prescription of atorvastatin (ATV) or simvastatin (SMV). Inclusion criteria were subjects of both genders between 21-99 years of age on either simvastatin or atorvastatin, but not both. Other inclusion criteria were adherence with 5 consecutive statin doses prior to sample collection and taking statins chronically for three months prior to enrolment. Exclusion criteria were patients unable or unwilling to give written consent and subjects who are pregnant or breast feeding.

Subjects were given 5 daily text message reminders and asked to record the time of drug ingestion at home (T_0_) on the evening prior to the study visit. Patients visited the clinic the morning after, and samples were collected in a fasting state at two time points T_1_ and T_2_, each separated by at least 3 hours. We obtained self-reported co-prescriptions from patient case report forms (CRFs).

#### Study Approval, Outcomes Linkage and Ethics

Under approval from Singapore Ministry of Health (MOH), National Registry of Diseases Office (NRDO) under request number Y21-S0002, 5-year clinical outcomes (MACE as a composite of Stroke, Acute Myocardial Infarction and All-Cause Mortality) up to June 2021 were obtained. The NRDO is a division within the Singapore Ministry of Health which collects data on major diseases and health conditions in Singapore, and is responsible for managing national registries in cancer, stroke, kidney failure and acute myocardial infarction. Access and use of the data was approved for use in this study by the NHG DSIRB Group D (2014/00856).

### Sample Analysis

#### LC-MS/MS Phenotyping

6 metabolites of Atorvastatin (ATV) metabolism and 2 metabolites of SMV metabolism were monitored in the LC-MS/MS assay (Supplementary Table S1, Supplementary Figure S1) using an Agilent 6495C.

An Agilent 6495C Mass Spectrometer was used to assay plasma samples. ATV is given as a medication to subjects in its active acid form, which can be metabolized into either atorvastatin lactone (ATVLAC), 2-hydroxy-atorvastatin (2OHATV) or 4- hydroxy-atorvastatin (4OHATV). The 2- and 4-hydroxy acid forms can also be metabolized to 2- and 4-hydroxy-atorvastatin lactone (2OHATVLAC/4OHATVLAC). In contrast, Simvastatin (SMV) is given in its lactone form, simvastatin lactone (SMVLAC)^8^. Detailed assay details, protocols and parameters are placed in Supplementary Methods and Supplementary Table S1.

Primary data was processed to optimize against environmental and technical noise by digital phenotyping (DPP) by accounting and controlling for LC-MS/MS batch effects and sensitivity drift, as well as variation in blood draw timing among different healthcare sites (Supplemental Methods, Supplemental Figures S2, 3).

#### Animal Study Design

All experiments were approved by the Biomedical Sciences Institute Singapore Institutional Animal Care Committee at A*STAR (Protocol No: 231805). Mice were maintained on a 12 h dark–light cycle, with ad libitum access to water and were fed with lipid-rich Western-Type Diet (D12079B, Research Diets, NJ).32 Male, 10-week C57BL/6J mice were purchased from INVIVOS Pte ltd, Singapore. All mice were administered an injection of 10^11^CFU of AAV8-mPCSK9-D377Y via tail vein to increase atherogenic LDL-C levels. The mice were fed a Western-type diet for a total of 4 weeks: 2 weeks before and 2 weeks after wire injury. Only male mice were used for this study to avoid the interference of estrogen effects on injury-related plaque with our target of interest. We performed wire injury^9^ on mice to accelerate atherosclerosis progression (Supplemental Figure S4). For the wire injury procedure, mice were anesthetized (100 mg/kg ketamine hydrochloride, 10 mg/kg xylazine i.p.) and subjected to endothelial denudation of the left common carotid artery using a 1 cm insertion of a flexible 0.36 mm guide wire through a transverse arteriotomy of the external carotid artery, as previously described^9^. Immediately prior to and for 2 days post-surgery, the analgesic buprenorphine (0.05–0.1 mg/kg) was administered via subcutaneous injection. Mice were randomly divided in four treatment groups: (Group 1 “S”) normal saline; (Group 2 “A”) Atorvastatin (16.66 mg/kg); (Group 3 “AO”) atorvastatin (16.66 mg/kg) + omeprazole (8.5 mg/kg) and (Group 4 “AL”) atorvastatin (16.66 mg/kg) + lansoprazole (8 mg/kg) (Supplemental Figure S5). All drugs and vehicle controls were administered by daily oral gavage, beginning on the day of wire injury and continuing for 2 weeks. The mice were euthanized at the end of treatment, plasma and the left and right carotid artery were collected for further analysis. The right carotid artery without wire injury served as matching control. 60 sections of 5 𝜇 m each was collected by microtome from the start of carotid bifurcation, such that 300𝜇m per mouse was collected. The first and last slides of each mouse, holding 6 cross sections each of equidistant tissue from each other were used for imaging and analysis of neointimal plaque and tunica media. Neointimal plaque area was normalized to media area to generate intima/media ratio as previously described^9^.

### Data Analysis

#### Analysis software

For DPP, metabolic-outcome and pharmacokinetic analysis, R v4.1.2 was the primary analysis software, including survival analysis, statistical tests, logistic regression, and cox hazards regression. We performed Log-Rank survival tests for populations separated by tertiles, while Cox Hazards Regression was used to generate all forest plots. Kaplan-Meier (KM) curves were used to visualize differences in survival. All global proportionality hazards assumptions were verified to hold true before reported. Benjamini-Hochberg correction was applied to statin metabolite hazards analysis and drug-drug interactions.

#### Multiple Testing

In survival analysis, Benjamini-Hochberg correction was performed on log-rank significance to control for multiple testing. For co-prescription analysis, Benjamini-Hochberg correction was applied to Mann-Whitney U-Test significance.

#### Missing Data and Imputation

Missing Data was not imputed.

#### Figure Statistics

Point graph error margins are calculated as tertiles from the median using the base R command *quantile(z, c(0.33, 0.66))*. All Boxplots display median and interquartile ranges. Error margins for forest plots correspond to 95% CI. P-values displayed on KM curves are Log-Rank P-values as calculated using the *survfit2* function of the *ggsurvfit* package.

### Resource Availability

#### Lead Contact

Further information and requests for resources and reagents should be directed to and will be fulfilled by the lead contact, Chester L Drum.

#### Data and Code Availability

The data that support the findings of this study are available from the corresponding author on reasonable request. Metabolic (.xlsx) and image (.tiff) data are deposited in the National University of Singapore private repository with ID 2014/00856 and UID *480f619vmfeb9wm5t834m*. The Institutional Review Board approved data sharing in compliance with ICH E6(R1) guidelines. A proposal with detailed aims, statistical plan, and other information/materials may be required to guarantee the rationality of requirement and the security of the data. The patient-level data, but without identifiers, will be shared after review and approval of the submitted proposal and any related requested materials. Outcomes data e.g. MACE can only be obtained from the NRDO via restricted access and approval and will require on-site supervision by NRDO personnel.

## Results

### Baseline characteristics of participants

1458 patients completed the study visits (CONSORT Diagram - Supplementary Figure S6). Complete clinical records were available for 1362 of these subjects (NUH, n=674; SGH, n=688). Clinical characteristics were similar in the 2 cohorts (Table 1). Overall, the mean age was 59.0±9.4 (range, 25-89) years. 1140 (84%) were men and 687 (50%), 322 (24%) and 336 (25%) were of Chinese, Malay and Indian ethnicity, respectively. As per the inclusion criteria, all patients were on chronic statin treatment upon enrolment. Doses of atorvastatin and simvastatin were identical at 40mg. Lipid profiles were similar between the two cohorts, with a median LDL-C of 2.2mmol/L (IQR=1.7-2.6) and 2.1mmol/L (IQR=1.7-2.8) at NUH and SGH, respectively (P=0.708). The average follow-up time for patients was 1388 days and differed for patients without MACE (1513 days), and those with MACE (722 days).

**Table 1.** Summary Statistics of Baseline Characteristics.

of patients in both NUH and SGH Cohorts.
| Variable | NUH Cohort<br>(n=674) | SGH Cohort<br>(n=688) | Full Cohort<br>(n=1362) | P-value |
| --- | --- | --- | --- | --- |
| **Baseline Characteristics** |  |  |  |  |
| **General** |  |  |  |  |
| Age | 57.9±8.6 | 60.1±10.0 | 59±9.4 | <0.001 |
| BMI | 27.5±5.2 | 26.5±4.5 | 27±4.9 | 0.002 |
| **Gender (n%)** |  |  |  |  |
| Male | 595 (88.3) | 544 (79.1) | 1139 (83.6) | <0.001 |
| Female | 79 (11.7) | 144 (20.9) | 223 (16.4) | <0.001 |
| **Race (n%)** |  |  |  |  |
| Indian | 164 (24.3) | 172 (25) | 336 (24.7) | 0.836 |
| Chinese | 297 (44.1) | 389 (56.6) | 686 (50.4) | <0.001 |
| Malay | 206 (30.6) | 116 (16.8) | 322 (23.6) | <0.001 |
| Others | 7 (1.0) | 11 (1.6) | 18 (1.3) | 0.506 |
| **Smoker (n%)** |  |  |  |  |
| No | 504 (74.8) | 594 (86.4) | 1098 (80.6) | <0.001 |
| Yes | 170 (25.2) | 94 (13.6) | 264 (19.4) | <0.001 |
| **Statin Profile (n%)** |  |  |  |  |
| **Statin Type (n%)** |  |  |  |  |
| Atorvastatin | 412 (61.1) | 564 (81.9) | 976 (71.6) | <0.001 |
| Simvastatin | 262 (38.9) | 124 (18.0) | 386 (28.3) | <0.001 |
| **Atorvastatin (mg)** |  |  |  |  |
| 5 | 4 (1.0%) | 6 (1.1%) | 10 (1.0%) | 1.000 |
| 10 | 27 (6.6%) | 30 (5.3%) | 57 (5.8%) | 0.500 |
| 15 | 0 (0%) | 1 (0.2%) | 1 (0.1%) | 1.000 |
| 20 | 79 (19.2%) | 89 (15.8%) | 168 (17.2%) | 0.193 |
| 30 | 2 (0.5%) | 2 (0.4%) | 4 (0.4%) | 1.000 |
| 40 | 284 (68.9%) | 342 (60.6%) | 626 (64.1%) | 0.009 |
| 50 | 0 (0%) | 1 (0.2%) | 1 (0.1%) | 1.000 |
| 60 | 3 (0.7%) | 16 (2.8%) | 19 (1.9%) | 0.034 |
| 80 | 13 (3.2%) | 77 (13.7%) | 90 (9.2%) | <0.001 |
| **Simvastatin (mg)** |  |  |  |  |
| 5 | 2 (0.5%) | 4 (0.7%) | 6 (0.6%) | 0.166 |
| 10 | 41 (10%) | 31 (5.5%) | 72 (7.4%) | 0.039 |
| 20 | 114 (27.7%) | 53 (9.4%) | 167 (17.1%) | 0.974 |
| 30 | 2 (0.5%) | 3 (0.5%) | 5 (0.5%) | 0.389 |
| 40 | 103 (25%) | 31 (5.5%) | 134 (13.7%) | 0.008 |
| 60 | 0 (0%) | 2 (0.4%) | 2 (0.2%) | 0.193 |
| **Medical History (n%)** |  |  |  |  |
| MI | 619 (91.8) | 602 (87.5) | 1221 (89.7) | 0.011 |
| Renal Disease | 96 (14.2) | 99 (14.4) | 195 (14.3) | 1.000 |
| Liver Disease | 21 (3.1) | 39 (5.7) | 60 (4.4) | 0.031 |
| Hypertension | 349 (51.8) | 479 (69.7) | 828 (60.8) | <0.001 |
| Diabetes Mellitus | 261 (38.7) | 326 (47.5) | 587 (43.1) | 0.001 |
| Hypercholesterolemia | 409 (60.7) | 547 (79.5) | 956 (70.2) | <0.001 |
| **Biochemical Measurements (median [IQR])** |  |  |  |  |
| **Renal Function** |  |  |  |  |
| Sodium (mmol/L) | 139.0 [137.0-140.0] | 138.0 [136.0-140.0] | 138.0 [137.0-140.0] | 0.052 |
| Potassium (mmol/L) | 4.3 [4.0-4.6] | 4.3 [4.0-4.6] | 4.3 [4.0-4.6] | 0.914 |
| Urea (mmol/L) | 5.3 [4.3-7.0] | 5.6 [4.4-7.7] | 5.4 [4.4-7.2] | 0.031 |
| Creatinine (μmol/L) | 88.0 [75.0-104.0] | 86.0 [72.0-106.0] | 87.0 [73.0-105.0] | 0.447 |
| eGFR (ml/min/1.73m<sup>2</sup>) | 81.8 [66.5-97.1] | 80.7 [61.2-97.5] | 81.4 [65.0-97.3] | 0.315 |
| **Lipid Profile** |  |  |  |  |
| Total Cholesterol (mmol/L) | 3.98 [3.5-4.5] | 3.92 [3.4-4.7] | 3.96 [3.4-4.6] | 0.971 |
| Triglycerides (mmol/L) | 1.39 [1.0-1.9] | 1.33 [0.9-1.9] | 1.37 [1.0-1.9] | 0.035 |
| HDL-C (mmol/L) | 1.06 [0.9-1.2] | 1.06 [0.9-1.2] | 1.06 [0.9-1.2] | 0.883 |
| LDL-C (mmol/L) | 2.2 [1.7-2.6] | 2.1 [1.7-2.8] | 2.2 [1.7-2.7] | 0.708 |
| Cholesterol / HDL Ratio | 3.7 [3.1-4.3] | 3.7 [3.1-4.5] | 3.7 [3.1-4.4] | 0.666 |
| **Liver Function** |  |  |  |  |
| AST (U/L) | 23.0 [19.0-26.0] | 31.0 [23.0-40.0] | 25.0 [20.0-31.0] | <0.001 |
| ALT (U/L) | 20.0 [16.0-27.0] | 26.0 [18.0-38.0] | 23.0 [17.0-32.0] | <0.001 |
| ALP (U/L) | 80.0 [64.0-99.0] | 82.0 [65.8-104.0] | 80.5 [65.0-101.0] | 0.290 |
| **Misc** |  |  |  |  |
| Creatine Kinase (U/L) | 132.0 [95.5-185.0] | 108.5 [77.0-174.2] | 126.0 [88.5-184.0] | 0.005 |
| **5-Year Clinical Outcomes (n%)** |  |  |  |  |
| MACE | 90 (13.4) | 143 (20.8) | 233 (17.1) | <0.001 |
| AMI | 39 (5.8) | 54 (7.8) | 93 (6.8) | 0.168 |
| Stroke | 16 (2.4) | 27 (3.9) | 43 (3.2) | 0.143 |
| All-cause mortality | 57 (8.5) | 95 (13.8) | 152 (11.2) | 0.003 |
| Missing Data | 3 (0.1) | 0 (0.0) | 3 (0.1) | - |

The recorded prevalence of ischemic heart disease (n=1221, 90%), hypertension (n=828, 61%), hyperlipidemia (n=956, 70%) and diabetes mellitus (n=587, 43%) was consistent with elevated cardiovascular risk. In total, 233 (17%) of patients experienced at least one MACE and 152 (11%) died. The percentage of patients who experienced MACE at NUH (n=90, 13.4%) versus SGH (n=143, 20.8%) was significantly different (χ2 p<0.001). The SGH cohort had a higher mortality rate than the NUH cohort [(n=95, 14%) vs (n=57, 8%), P=0.003].

### Co-prescriptions and their effects on statin lactonization and clinical outcomes

Current DDI’s include fibrates, antifungal drugs, immunosuppressives and dietary substances such as grapefruit which can result in reduced effectiveness and side effects^10^. We obtained self-reported co-prescriptions from patients on their first visit (Table 2) and performed multiple hypotheses testing for differential expression analysis of plasma ATVLAC on all patients. 776 subjects on ATV had full co-prescriptions data, of which 774 had complete outcomes data. 51 unique co-prescriptions were recorded across all patients. Aspirin had the highest co-prescription rate (67%), followed by omeprazole (54%) and bisoprolol (45%) (Table 2).

**Table 2.**
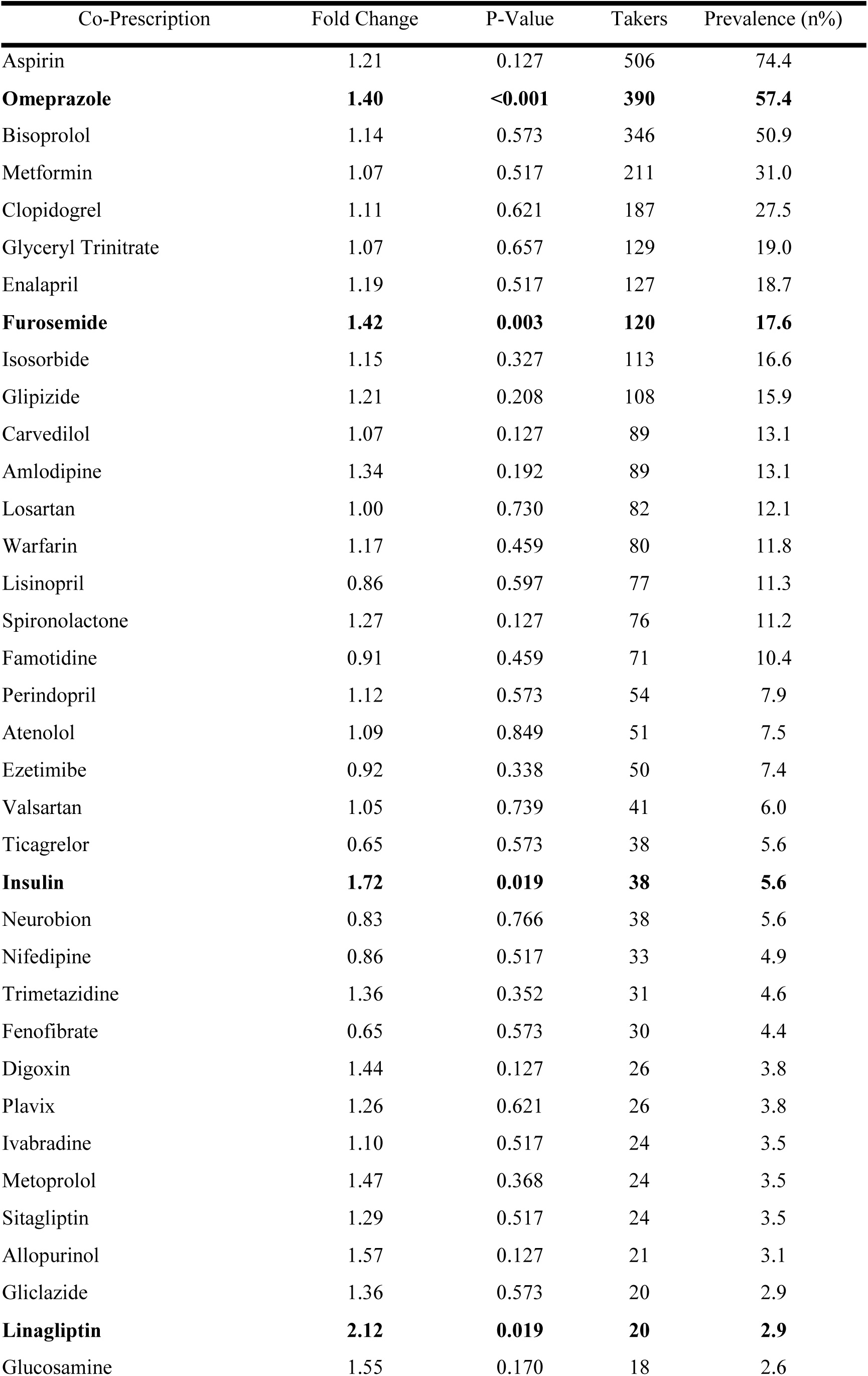
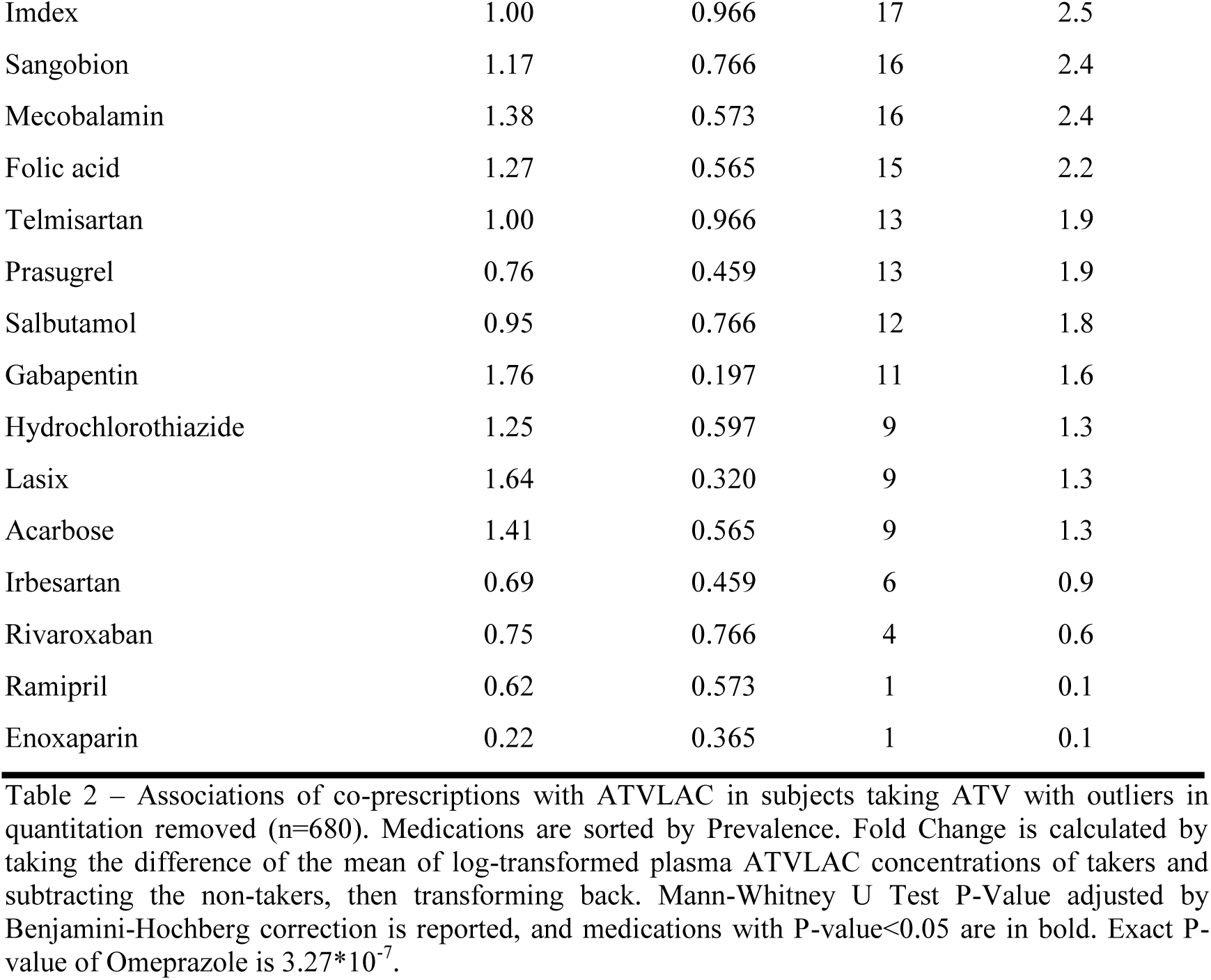
Association of co-prescriptions and ATVLAC.

After Benjamini-Hochberg correction, insulin (P=0.019), linagliptin (P=0.019), omeprazole (P<0.001) and furosemide (P=0.002) were significantly associated with elevated atorvastatin lactone (Figure 1a, Table 2). Diabetes mellitus reduces the clearance of ATVLAC^11^. Thus, the association between insulin, linagliptin and plasma ATVLAC is expected. However, the association between omeprazole and elevated ATVLAC was unexpected. Omeprazole, the formulary offering for PPI’s in NUH and SGH, was the second most prescribed medication at 54% overall and 57% within subjects taking ATV. Patients taking omeprazole had the most significant elevation in plasma ATVLAC (1.41-fold, P<0.001) (Figures 1a, b) among all other drugs. In addition, these patients had higher rates of MACE (HR=1.52, CI=1.04–2.21, P=0.028) (Figure 1c).

**Fig. 1.**
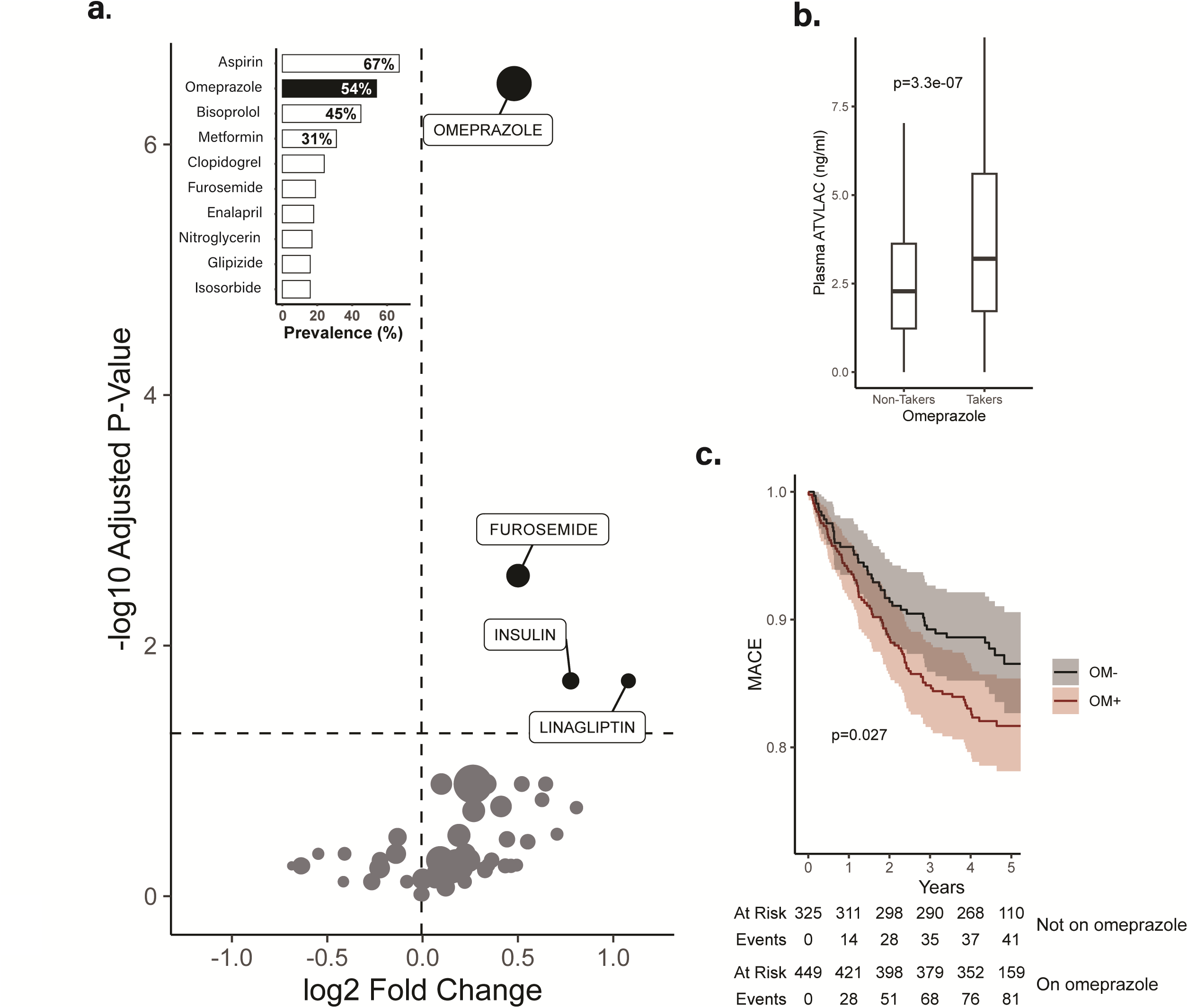
Drug-drug interactions and plasma statin lactone. **a)** Volcano plot of ATVLAC and co-prescribed oral medications (point area proportional to prevalence). Y-axis displays adjusted log_10_ Mann-Whitney U-Test p-value adjusted for multiple testing, x-axis displays the log_2_ fold-change. Dashed lines demarcate a neutral fold-change of 0, and a p-value cut-off of 0.05. Inset: Co-prescription frequencies in the general cohort, **b)** Plasma ATVLAC of participants on and off omeprazole co-prescription, Mann-Whitney p-value displayed, **c)** KM Survival Curves of the ATV Cohort, segmented by Omeprazole (OM-, OM+) co-prescription, log-rank p-value displayed. Similar trends were observed even in patients who were not on concurrent clopidogrel prescription alongside statins and omeprazole (Supplemental Figure S7). OM+/-: On or off omeprazole co-prescription.

Simvastatin is manufactured and administered in lactone form. In patients on simvastatin, none of the co-prescribed medications were associated with either elevated SMVLAC or MACE (Supplementary Table S2, Supplemental Discussion).

### Association of omeprazole with MACE is independent of clopidogrel and Framingham Risk Factors

A PPI drug interaction which decreases the activity of clopidogrel has previously been suggested as a cause of poor outcomes in PPI cardiovascular patients^12,13^. We thus performed the same analysis on patients who took omeprazole but not clopidogrel and found similar results for MACE (HR=1.69, CI=1.09–2.61, P=0.018) (Supplemental Figure S7a). In addition, the association of omeprazole with increased rates of MACE was preserved while controlling for Framingham risk factors (HR=1.65, CI=1.11–2.45, P=0.013) (Figure 1c, e, Supplementary Figure S7b).

### Plasma atorvastatin lactone is associated with clinical outcomes

We found pair-wise associations (P<0.05) of all statin metabolites with 5-year MACE and all-cause mortality in both NUH and SGH cohorts. In a ranking of all metabolites, ATVLAC (HR=1.30, CI=1.16–1.45, P<0.001) and 4OHATVLAC (HR=1.30, CI=1.20–1.40, p<0.001) (Figure S8) had the largest hazard ratio. After correcting for multiple testing, ATVLAC was the only metabolite significant in both cohorts (NUH, P=0.006, SGH, P=0.009, combined, P<0.001). In a Kaplan-Meier (KM) analysis of each metabolite segmented into tertiles, ATVLAC was also the strongest predictor of MACE in each (Supplemental Figure S9a, b) and both cohorts (Figure 2a, P<0.001). We observed the same trend of plasma SMVLAC in patients on SMV (Supplemental Results).

**Fig. 2.**
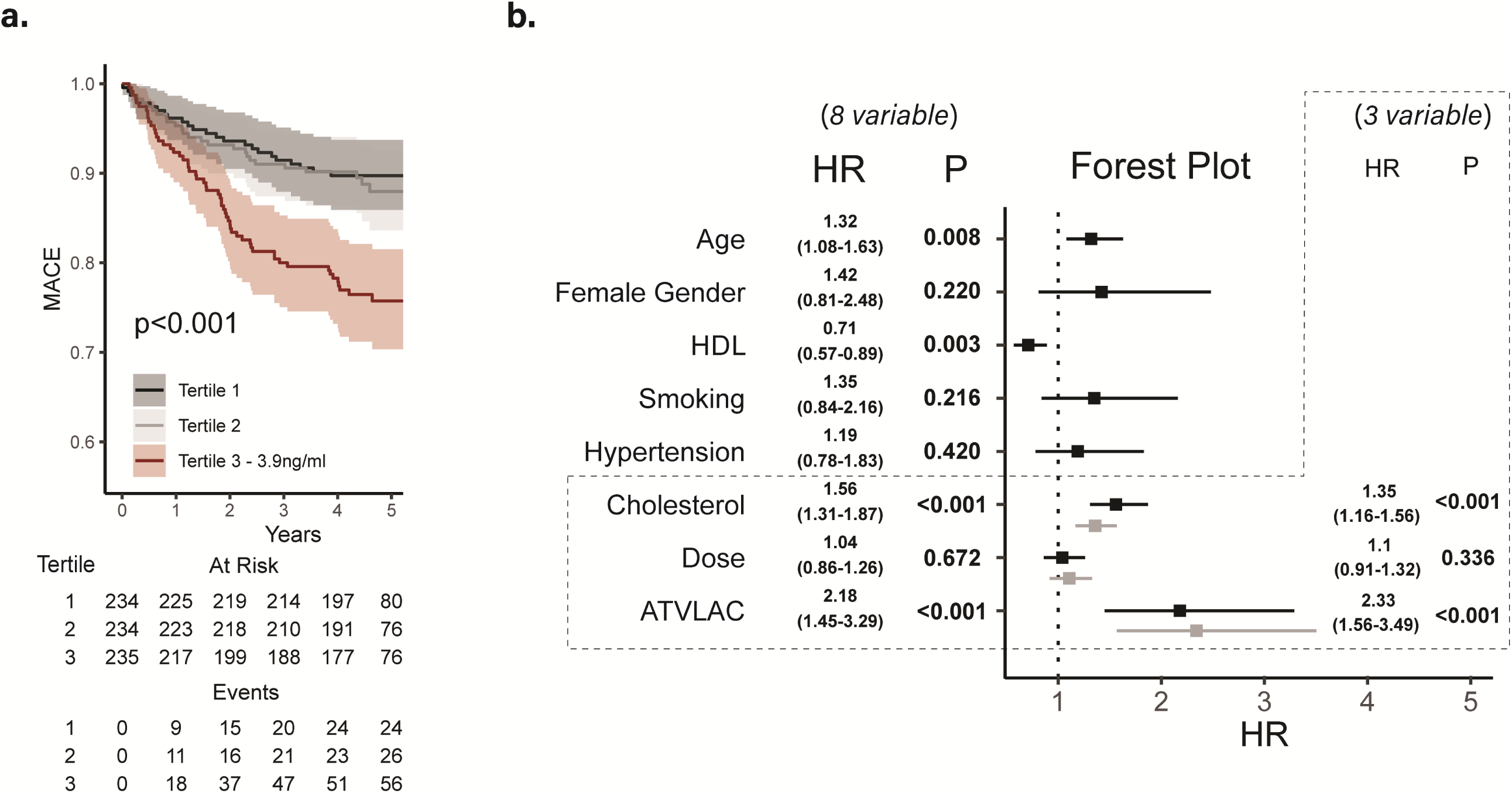
Clinical outcomes, ATVLAC and independence of findings from LDL-C. **a)** Kaplan-Meier curve, with log-rank p-value displayed, for plasma ATVLAC tertiles against 5-year MACE in the combined cohort, number-at-risk table displayed, **b)** Multivariable Cox proportional hazards model including Framingham risk factors and ATVLAC≥3.9ng/ml. Boxed portion indicates a three variable model of total cholesterol, statin dose and ATVLAC.

To determine a threshold of statin lactone concentration, we examined the Kaplan-Meier survival curves under tertiles, and observed the third tertile of ATVLAC ≥3.9ng/ml as a threshold of risk for both MACE (HR=2.45, CI=1.60–3.44, P<0.001) and all-cause mortality (HR=3.18, CI=1.96–5.16, P<0.001) (Supplementary Table S3) in the combined cohort.

### Independent association of ATVLAC with adverse outcomes

To determine if the association was independent of LDL-C levels, we segmented patients by tertiles of achieved LDL-C. Within these tertiles [T1 (<1.84mmol/L); T2 (1.84mmol/L to 2.44mmol/L); T3 (≥2.44mmol/L)], we found no evidence for correlation between ATVLAC and LDL-C [T1, ρ=0.02, P=0.775; T2, ρ=0.05, P=0.500; T3, ρ=-0.07, P=0.336] (Supplementary Table S4). Likewise, we tested the association of outcomes with ATVLAC ≥3.9ng/mL segmented to tertiles of LDL-C and found more events at all levels of LDL-C [T1 (HR=5.77, CI=2.33–14.3, P<0.001); T2 (HR=2.06, CI=0.97–4.38, P=0.061); T3 (HR=1.74, CI=1.00–3.06, P=0.049)] (Supplemental Figure S9c, d, e).

To investigate the possible confounding effect of dosage (higher doses may correlate with increased risk), we analyzed subjects only taking 40mg/day ATV (n=454, MACE events=70). ATVLAC ≥3.9ng/ml continued to predict both MACE (KM, P<0.001; HR=2.46, CI=1.53–3.96, P<0.001) and all-cause mortality (HR=3.22, CI=1.72–6.02, P<0.001). Normalizing ATVLAC to drug dose similarly predicted MACE (HR=1.86, CI=1.27–2.71, P=0.001) and all-cause mortality (HR=2.02, CI=1.25–3.27, P=0.004) (Supplementary Figures S9f, g).

To adjust for potential bias, we performed a Cox Hazards multivariable analysis. As LDL-C and total cholesterol are well-known drivers of cardiovascular disease, in a 3- variable model including dose with either total cholesterol or LDL-C, ATVLAC ≥3.9ng/ml demonstrated the strongest and most statistically robust hazard for adverse outcome (HR=2.33, CI=1.56–3.49, P<0.001, HR=2.36, CI=1.56–3.56, P<0.001, respectively) (Figure 2b) When the Framingham variables sex, age, HDL-C, smoking status, hypertension, total cholesterol, and drug dose were included, ATVLAC ≥3.9ng/mL remained a strong independent predictor of MACE (HR=2.18, CI=1.45– 3.29, P<0.001) and all-cause mortality (HR=2.58, CI=1.53–4.37, P<0.001) (Figure 2b). When effective glomerular filtration rate (eGFR) was included as a variable addressing renal function, ATVLAC ≥3.9ng/ml remained significant in predicting increased hazard (HR=1.85, CI=1.24-2.75, P=0.003, Supplementary Table S5).

We validated the risk threshold across institutions by computing the optimal diagnostic threshold using the Area Under Receiver Operating Curve (AUROC). Defining risk as the presence of a 5-year event, ATVLAC of 4.1ng/mL was found to be the optimal threshold, similar to that identified by the Kaplan-Meier survival analysis for tertiles (Supplementary Figure S10). Compared to the ROC constructed with Framingham risk variables, a logistic model constructed with ATVLAC and LDL-C demonstrated comparable AUC in the NUH, SGH, and combined cohort (AUROC = 0.70 vs 0.70, 0.69 vs 0.67, and 0.70 vs 0.69 respectively).

### Interactions between ATVLAC tertile threshold and omeprazole co-prescription

We hypothesized that if the event rates associated with omeprazole were due to an increase of ATVLAC above 3.9ng/mL, omeprazole would not be predictive at ATVLAC levels below 3.9ng/mL. Remarkably, in low-risk patients with ATVLAC <3.9ng/mL (n=453, events=50) omeprazole was not associated (HR=1.00, CI=0.57–1.74, P=0.992) whereas in high-risk patients with ATVLAC ≥3.9ng/ml (n=227, events=57) (Supplementary Figures S7c, d), omeprazole remained predictive (HR=2.70, CI=1.27–5.70, P=0.009). Lastly, we examined ATVLAC as a function of co-prescribed drugs and observed higher levels ATVLAC at all levels of polypharmacy in the presence of omeprazole (Supplementary Figure S9e).

### Omeprazole co-administration increases atorvastatin lactonization in mice

To validate our findings, we initiated a 4-week interventional mouse study (n=32), simulating the increased ATV lactonization using omeprazole co-prescription. As omeprazole is the formulary offering in NUH and SGH for PPI’s, we also validated our results using another proton pump inhibitor, lansoprazole. All mice were fed western type diet for 4 weeks and injected with AAV8-*PCSK9*-D377Y to increase circulatory LDL-C levels at the start of western-diet feeding. At the end of the second week of feeding, the negative control arm was given saline (S arm), while the positive control arm was given atorvastatin (A Arm). The omeprazole arm was co-administered omeprazole alongside atorvastatin (AO Arm), and the lansoprazole arm was co-administered lansoprazole alongside atorvastatin (AL Arm) (Supplemental Figure 4). To interrogate endothelial plaque formation, we performed wire injury-induced de-endothelialization of the left common carotid artery in all mice at the end of the second week (Supplemental Figure 5). Mice were sacrificed at the end of 4 weeks of feeding, and plasma was collected for LC-MS/MS analysis. Carotid artery sections were also collected, embedded in paraffin, and sectioned for neointimal lesion quantification (see Supplemental Methods and Supplemental Results).

As expected, mice in the negative control group had no plasma ATV metabolites (Supplemental Figure S11, Supplementary Table S6a). In contrast, mice in both the omeprazole and lansoprazole arms had significantly higher plasma ATVLAC compared to the positive control arm normalized to ATV, with mice in the lansoprazole arm exhibiting the highest lactonization (AO v A: 1.87-fold, p=0.004, AL v A: 3.56-fold, p<0.001, Figure 3b, Supplementary Table S6b).

**Fig. 3.**
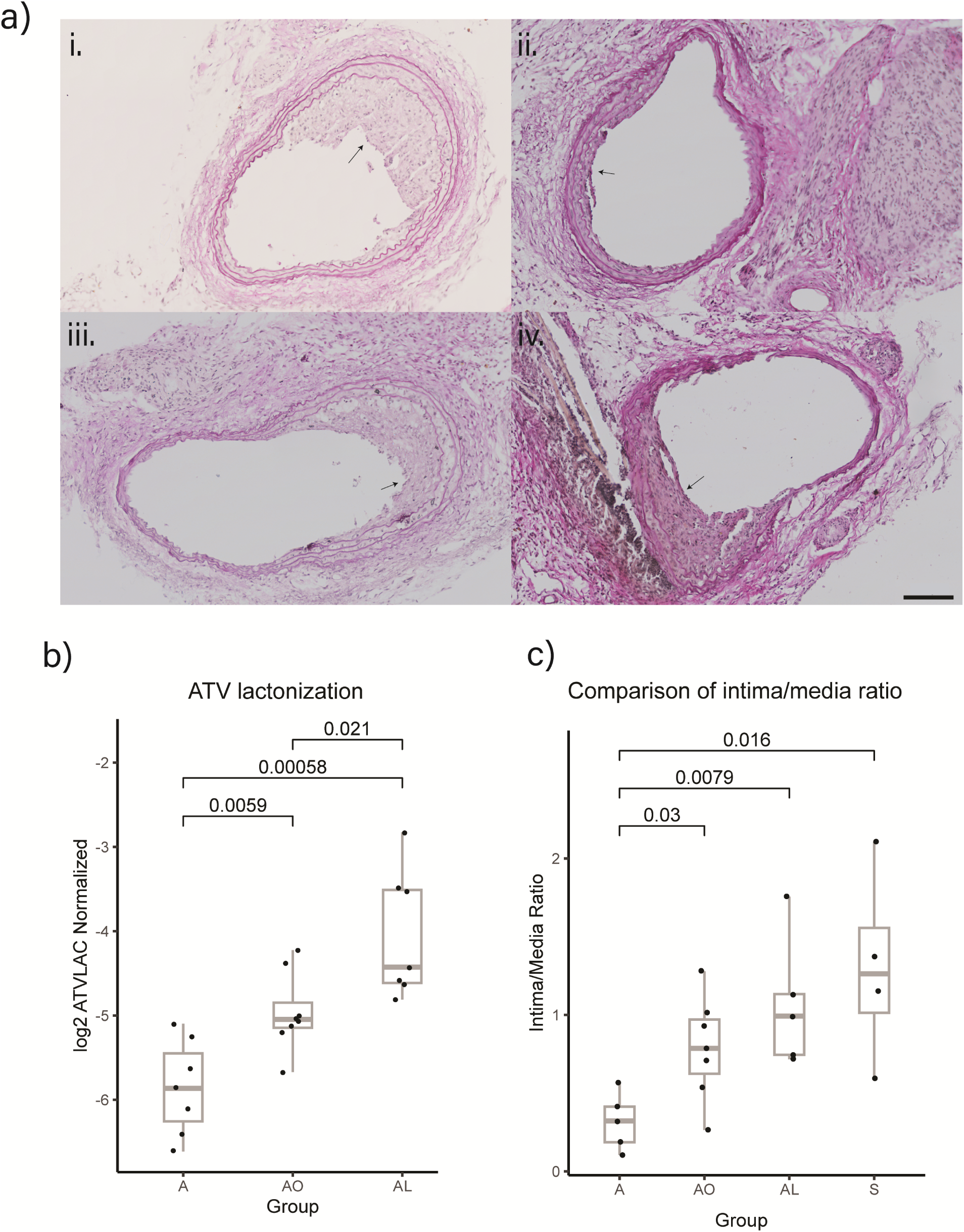
Wire Injury Mice Model of ATVLAC-mediated neointima formation. **a)** Representative carotid sections from: i. S, ii. A, iii. AO, and iv. AL arms. Arrows indicate neointima, 100µm scale bar on bottom right. **b)** Comparison of ATV lactonization for arms A, AO and AL (n=26). **c)** Comparison of intima/me­ dia ratio, a measure of plaque formation, of arms A (n=5), AO (n=7), AL (n=5) and S (n=4). Mann-whitney U test p-value displayed. See supplemental tables S6, S7 for all model and test statistics. A, ATV: atorvastatin, AO: atorvastatin with omeprazole, AL: atorvastatin with lansoprazole, S: saline.

### PPI increases neointima-to-media formation, associated with ATV lactonization

We next assessed neointimal plaque formation in the mice. As expected, the saline arm had the highest neointimal plaque area (mean = 23.4 ± 14.7%), while the atorvastatin arm had the lowest plaque area (mean = 5.6 ± 4.3%). Interestingly, the PPI arms showed significantly higher neointimal plaque area (AO: 15.7 ± 4.9 %, AL: 19.2% ± 8.0%) compared to the atorvastatin arm (AO: 2.97-fold, p=0.018, AL: 3.84- fold, p=0.016, Figure 3c, Supplementary Table S6c), suggesting that co-administration of PPIs with atorvastatin reduced the atheroprotective effects of ATV. In addition, plasma lactone levels were significantly associated with neointimal plaque area (Spearman’s 𝜌 = 0.66, p=0.0046, Figure S12), suggesting that the increased lactonization directly leads to increased plaque formation.

To account for section distance from the injury, we constructed a linear one-hot encoded multivariable model, controlling for distance of each individual section from the carotid bifurcation (Supplementary Table S7a). With the A arm as reference, we likewise observed significant differences in the S, AO and AL groups, whereby co-administration of omeprazole led to a 0.43 (CI: 0.18-0.68, P<0.001) increase in intima/media ratio. Co-administration of lansoprazole led to a 0.69 (CI: 0.44-0.96, P<0.001) increase in intima/media ratio, compared to the reference A arm. To confirm class effect, we tested lansoprazole co-administration and found higher plaque formation compared to omeprazole. The co-administration of lansoprazole resulted in significantly increased neointimal plaque area (𝛽 =0.27, CI=0.08-0.46, P=0.005, Supplementary Table S7b).

## Discussion

We conducted a prospective, multi-centre, observational 5-year cardiovascular outcomes study to examine the relationship between statin metabolism, clinical risk factors and common co-prescriptions. We discovered a novel drug-drug interaction between two of the most commonly co-prescribed drug classes: statins and PPIs. The PPI omeprazole induces dysmetabolism of the active form of statin drug to produce hydrophobic, statin lactones, a form of the drug known to be toxic from prior studies. The use of omeprazole was associated with 5-year MACE. We validated our observations in a mouse model of atherosclerosis and observed both elevated statin lactonization and increased neointimal plaque formation with co-administered PPIs.

Statins are multi-ring molecules which use a dihydroxyheptanoic acid moiety to competitively inhibit the cholesterol synthesis enzyme, HMG Co-A reductase. Following hepatic metabolism, the acid side chain can form a lactone ring, resulting in metabolites which are ∼1000-fold more hydrophobic than the acid form, cross cell membranes passively and do not inhibit HMG CoA reductase^14^. Statin lactones have previously been associated with statin toxicity, and reduced metabolic efficiency, mediated by reduced mitochondrial activity and inhibition of the Q_o_ site of the CIII respiratory complex^5,15^.

Our observation that omeprazole co-prescription was associated with increased 5-year MACE and elevated statin lactone concentrations is important because co-prescription of omeprazole is common, and prior observational studies have found an association between omeprazole and adverse cardiovascular outcomes^16–18^ without a clear explanatory mechanism. The risk of increased MACE is easily modifiable with conversion to an alternative antacid, and we found no evidence for increased lactone levels with H2 receptor blockers. Omeprazole is a CYP3A4 inhibitor^19^, a known inducer of UGT1A activity^7,20^ and has been proposed to decrease the effectiveness of clopidogrel, which requires CYP3A4 for bioactivation^21^. We ruled out a clopidogrel- dependent effect by repeating our findings in patients on omeprazole and statin but not clopidogrel.

In our study, omeprazole is a clear outlier in its strength of association with respect to statin lactone concentrations (Figure 1a), an effect independent of polypharmacy. We note that while omeprazole is highly predictive of outcome in patients with ATVLAC ≥3.9ng/ml, there is a complete lack of association with outcome in patients with ATVLAC <3.9ng/mL. Whereas ATV is administered in an acid form and subsequently converted to lactone, SMV is administered as a lactone prodrug and rapidly converted to the acid form. Thus, in our study, SMV serves as a negative control for lactonization and, indeed, we observe that patients taking SMV are not affected by omeprazole co-prescription.

We identified a threshold concentration of ATVLAC (>3.9ng/ml) as highly predictive of increased MACE when measured 13 hours post-dose, a timing amenable to fasting phlebotomy performed for outpatient cholesterol monitoring. Plasma statin levels are correlated to their dose, which may be increased in response to cardiovascular risk, thus it was critical to demonstrate that the observed effect of statin lactonization on MACE was independent of Framingham risk variables, statin dose and achieved LDL-C levels. In patients taking a uniform dose of statin, statin lactone levels remained predictive of outcome, and we found no correlation between statin lactone levels and achieved LDL-C. The prognostic ability of lactones was observed at all LDL-C levels, and most interestingly, in patients who achieved low LDL-C [<1.84mmol/L] while on statin treatment (Supplemental Figure S9c). Lastly, in multivariable models that included Framingham risk factors, achieved LDL-C and statin dose, statin lactones again remained independently predictive of MACE outcomes and exhibited the largest independent hazard ratio of any variable in the model. To examine a class effect, we found similar findings with SMV at a threshold of SMVLAC (>0.16ng/ml).

The causative effect of omeprazole co-administration on ATV lactonization and cardiovascular risk was examined using a mouse model in which vessel de-endothelialization was induced by wire injury. This interrogates vascular remodelling and accelerated neointimal plaque formation via endothelial damage. After observing increased lactonization and accelerated plaque progression with PPI co-prescription, we hypothesize that the increase in plaque formation we observed may have been driven by the inhibition of autophagy and accumulation of oxidative stress. ATV lactone mediates mitochondrial inhibition^5^, and its previously identified inhibition of respiratory complex CIII results in the formation of Reactive Oxygen Species (ROS)^22^. CIII function is crucial for autophagy^23^, and the inhibition of autophagy has been shown to lead to increased plaque formation^24,25^. Interestingly, lansoprazole was a more potent inducer of ATV lactonization, with significantly increased neointimal plaque formation compared to omeprazole. This observation is consistent with the higher bioavailability of lansoprazole compared to omeprazole^26^.

Our study has several limitations. First, to create a scalable protocol for recruiting large numbers of participants, measurements were taken in the late elimination phase of the pharmacokinetic absorption curve, meaning we did not perform a 24-hour complete PK study on every study subject. Nevertheless, our separate intra- and inter-individual validation study demonstrated acceptable predictive consistency of this sampling regimen, supporting its broader use. Also, for the purpose of patient inclusion, the time of drug ingestion was self-reported which likely introduced variability into the PK model, we addressed this potential source of variability by testing and validating a digital phenotyping protocol further described in the methods. Third, unmeasured environmental and dietary factors including objective measures of statin adherence over the 5-year follow-up period were not possible within the scope of the project design.

This study describes the discovery of a novel drug-drug interaction between statins and proton pump inhibitors, two of the most commonly prescribed drug classes in the world, through the use of real-world pharmacokinetic phenotyping. Statins are a cornerstone of cardiovascular risk reduction and the potential induction of statin dysmetabolism via common PPI co-prescriptions deserves further study.

## Author Contributions

Conceptualization: EGCH, CLD, MG, RRS, EAL

Study design: CLD, TES, JT

Methodology (metabolomic): EGCH, CLD, LP

Methodology (genomic): EGCH, WLH, YJL, FA, LJJ

Methodology (animal): EGCH, MKS, RRS, EAL

Investigation (metabolomic): EGCH, CLD, WLH, LP

Investigation (animal): EGCH, LXY

Visualization (metabolomic): EGCH, WLH

Visualization (animal): EGCH

Funding acquisition: CLD, TES, JJ

Project administration: EGCH, CLD, NML

Supervision: CLD

Writing – original draft: EGCH, CLD, WLH, LP

Writing – review & editing: all

All authors have approved the final version of this paper.

## Data Availability

The data that support the findings of this study are available from the corresponding author on reasonable request. Metabolic (.xlsx) and image (.tiff) data are deposited in the National University of Singapore private repository with ID 2014/00856 and UID 480f619vmfeb9wm5t834m. The Institutional Review Board approved data sharing in compliance with ICH E6(R1) guidelines. A proposal with detailed aims, statistical plan, and other information/materials may be required to guarantee the rationality of requirement and the security of the data. The patient-level data, but without identifiers, will be shared after review and approval of the submitted proposal and any related requested materials. Outcomes data e.g. MACE can only be obtained from the NRDO via restricted access and approval and will require on-site supervision by NRDO personnel.

## Acknowledgments

This work was supported by the National Medical Research Council (CSAINV17nov012), ASTAR Strategic Positioning Fund (SPF2014/001), ASTAR Industrial Alignment Fund (IAF-PP) and funding from the Economic Development Board-IPP with Agilent Technologies. We thank Prof. Mark Richards and Prof. Howard Bauchner for critical reading of the manuscript.

## Trial Registration

The study is registered at ClinicalTrials.gov (https://clinicaltrials.gov/study/NCT03042286).

## Declaration of Interests

The authors declare no competing interests.

## Supplemental Information

Supplemental Methods

Supplemental Results

Supplemental Discussion

Figures S1-15

Tables S1-9

References 27-38

## Notes

### Competing Interest Statement

The authors have declared no competing interest.

### Clinical Trial

NCT03042286

### Author Declarations

The Institutional Review Board approved data sharing in compliance with ICH E6(R1) guidelines. Access and use of the data was approved for use in this study by the NHG DSIRB Group D (2014/00856).

